# Modeling brings additional insights into the kinetics of SARS-CoV-2 neutralizing antibody

**DOI:** 10.1101/2021.10.13.21264693

**Authors:** Prague Mélanie, Quentin Clairon, Jérémie Guedj, Rodolphe Thiébaut

## Abstract

Following the paper by Seow et al. published in Nature Microbiology, we reanalyzed the publicly available data using dynamical models of humoral response. The main conclusion is that, from available data, we can demonstrate that the decline of neutralizing antibodies (as measured with ID50) is biphasic, which is compatible with two types of antibody secreting cells (short lived and long lived). We found that lower bound of half life of the long-lived antibody secreting cells is 450 days. Moreover, our model predicts that the neutralizing antibody response could be more durable than suggested (up to 129 days for individuals with no requirement of supplemental oxygen and up to 175 days for others). A result which adds insight on the longevity of immune response.

---

We have read with interest the paper by Seow et al. [4] that offers a unique description of the quantitative and functional kinetics of SARS-CoV-2 antibody. The authors convincingly demonstrate that the magnitude of the neutralizing antibody response is associated to disease severity, as defined by the need for supplemental oxygen during infection. Furthermore, this response is characterized by a high variability across individuals, with some individuals showing a modest neutralizing antibody titers and some others with detectable levels up to 50 days after symptom onset. We here would like to complement their findings by a mathematical and statistical modeling of their data, which offers additional insights into the potential duration of the response and the mechanisms inherent to antibody kinetics.

We reanalyzed the repeated measures of the neutralizing antibody titres, *ID*_50_(*t*), using a model of antibody expansion and contraction previously developed in the context of Ebola viral infection [2]. Briefly, the model considers two populations of antibodies secreting cells (ASCs), the short-lived (*S*, having a half-life noted 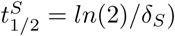 and the long-lived (*L*, having a half-life noted 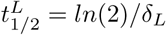). Assuming that the neutralizing function is proportional to the number of circulating functional antibodies, the kinetics of antibody can then be written as 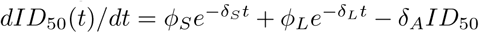, in which *ϕ*_*S*_ and *ϕ*_*L*_ represent the antibodies influx from short-lived and long-lived ASCs and *δ*_*A*_ is the antibodies decay rate. Next we used a mixed-effect model, a statistical approach that borrows strength from the whole sample [1] to estimate parameters using the SAEM algorithm implemented in Monolix [3], to estimate the parameters of the model, their distribution in the population and the impact of disease severity on the model parameters.

The kinetics of the neutralizing antibody in the 83 individuals presented in Seow et al. [4] could be well captured by the model. The model predicted a peak of antibodies that was influenced by the disease severity through the antibodies influx from short-lives ASC, *ϕ*_*S*_ is increased by 14 [4; 48] change fold in individuals with severe symptoms (*p <* 0.001). After the peak, the antibodies declined exponentially with no differences in ASCs half-lives between severe and non-severe individuals, which was attributed to the decline of short lived cells with a half-life of 4.8 [2.0; 11.9] days. In a second phase, the model predicted a slower decline of antibodies, revealing a smaller but steady contribution to antibody production due to long lived cells. Although the half-life of longlived cells could not be precisely estimated, determination by profile likelihood revealed that half-life values were larger than 450 days. Importantly, a simplified model neglecting the contribution of long lived cells significantly deteriorated data fitting (P=0.001, Likelihood Ratio Test), supporting the existence of two distincts compartments of antibody producing cells with different half-lives. The bi-phasic trajectories for average severe and non-severe individuals are depicted in Figure 1 panel A.

**Figure 1:**
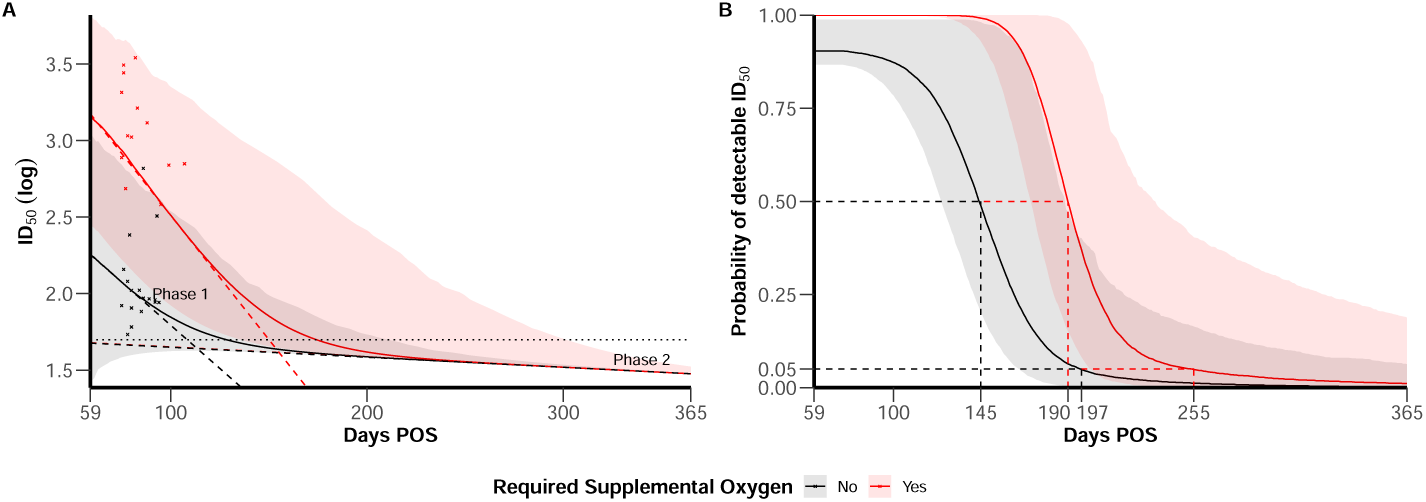
(A) *ID*_50_ trajectories for individual with mean characteristics according to disease severity (severe in red vs. non-severe in black), 95% confidence bands account inter-individual variability. Horizontal line represents the limit of detection at *ID*_50_ = 50. Dashed line materialise the slope of the bi-phasic decline of *ID*_50_. Red and black crosses represent available data after 59 days post symptom onset (POS) in the dataset for severe and non-severe individuals. (B) Probability of detectable *ID*_50_ along time post symptom onset according to disease severity (severe in red vs. non-severe in black). Plain line represent the median behavior and 95% confidence bands account for both uncertainty in parameters estimation and inter-individual variability.

Using the estimated model parameters, we next aimed to predict the longevity of the response, defined as the time to reach undetectable neutralizing antibody (*ID*_50_ *<* 50). The probability of detectable viral load each day post symptom onset is depicted in Figure 1 panel B. Consistent with the original findings, we estimated that the protection duration was larger in individual with severe disease. However, the estimated impact of long-lived cells led us to longer period of protection than initially described, with mean values of 175 and 129 days post symptom onset in individuals with severe and non-severe symptoms, respectively. This mean estimates nonetheless masks a large between individuals variability, with prediction intervals ranging from 136 to 302 days in individuals with severe symptoms, compared to 0 to 211 in individuals with non-severe symptoms.

By modelling the biphasic decline of antibody levels reported in Seow et al., this work is revealing the contribution of long-lived antibody producing cells. Although the peak of antibody is larger in individual with severe symptoms, our analysis did not find significant differences in the half-life of the short- and long-lived ASCs according to disease severity. Hence, the differences observed according to disease severity stand during the acute phase and do not seem to compromise the contribution of long-lived antibody producing cells. As a consequence of this biphasic decline, our modeling analysis of the data presented in Seow et al. predicts that the neutralizing antibody response could be more durable than suggested. We encourage collection of the kinetics of functional antibody over time in larger populations, which will be critical to refine these estimates and to evaluate other sources of variability in the population. These analyses will also need to be complemented by a deeper characterization of the cells that contribute to short- and long-term antibody kinetics.

## Data Availability

Data used are available in the supplementary material of this paper: https://www.nature.com/articles/s41564-020-00813-8

## Notes

### Competing Interest Statement

The authors have declared no competing interest.

### Funding Statement

This study did not receive any funding

### Author Declarations

The data are available as a supplementary information of this paper: https://www.nature.com/articles/s41564-020-00813-8

